# Systematic Review: Recommendations for Rehabilitation in ASD and ID from Clinical Practice Guidelines

**DOI:** 10.1101/2021.04.05.21254892

**Authors:** Jordan Wickstrom, Kristin Dell’Armo, Emma Salzman, Jessica L. Hooker, Abigail Delehanty, Somer Bishop, Marc J. Tassé, Amy M. Wetherby, Antonia M. H. Piergies, Diane Damiano, Alexandra Rauch, Audrey Thurm

**Author notes:** **Corresponding Author**: Audrey Thurm, PhD is the corresponding author, and she can be contacted at the address Magnuson Clinical Center, Room 1C250, MSC 1255, Bethesda, MD 20814, via phone at or email at. Reprints may also be obtained from Dr. Thurm. **Presentation of Material**: This material has not been presented anywhere. **Conflicts of Interest**: None.

## Abstract

**Objective:** Autism spectrum disorder (ASD) and intellectual disability (ID) were selected for inclusion in the development of a Package of Interventions for Rehabilitation for the World Health Organization (WHO). Here, we describe results of a search of guidelines for these conditions.

**Methods:** A literature search for clinical practice guidelines was performed for ASD and ID. Using the Appraisal of Guidelines for Research and Evaluation instrument and other inclusion and exclusion criteria, guidelines were identified for abstraction of recommendations, which were collated into categories based on diagnosis, patient age, type and target of service, valence, and level of evidence.

**Results:** Six guidelines remained after screening. Four ASD guidelines focused on diagnosis, management and support, and two ID guidelines covered the prevention, assessment, and management of challenging behavior and mental health problems, with 386 and 138 recommendations resulting for each group, respectively. Out of 524 total recommendations, 212 ASD and 58 ID recommendations focused on intervention. The primary type of intervention pertained to pharmacology for each group (27% ASD, 29% ID), followed by psychosocial (21%) and biomedical (23%) for ASD and behavioral (14%) and psychological (14%) for ID. Regarding target of intervention, recommendations primarily focused on coexisting conditions for each group (56% ASD, 93% ID) and core symptoms for ASD (26%), whereas adaptive functioning received little attention (11% ASD, 7% ID).

**Conclusions:** Although these six guidelines provided many recommendations for comorbid and specific problems in ASD and ID, very few recommendations targeted core symptoms, and many were based on expert opinion. A vital next step includes identifying relevant interventions from the guidelines or that are missing to be included in the WHO Package of Interventions for Rehabilitation.

---

The World Health Organization (WHO) launched the Rehabilitation 2030 Call for Action initiative to call attention to and strengthen rehabilitation in health systems worldwide.^1-3^ Rehabilitation is among the primary services included in universal health coverage and is defined as a set of interventions designed to optimize functioning and reduce disability in individuals with health conditions in interaction with their environment.^3^ The WHO Rehabilitation Programme is developing a Package of Interventions for Rehabilitation aimed at the national and sub-national levels^1^ with a specific focus on low-and middle-income countries. To achieve this goal, the Package of Interventions for Rehabilitation will provide information on which evidence-based interventions to provide, how to perform such interventions, and associated resource requirements relevant to rehabilitation that will help countries plan, budget, and integrate interventions across all service delivery platforms.

The development of the Package of Interventions for Rehabilitation follows a stepwise approach.^1^ Twenty health conditions were initially selected based on prevalence, associated level of disability (using disability weights),^4^ and expert opinion. The health conditions outlined for the development of the Package of Interventions for Rehabilitation are congenital or acquired conditions, and non-communicable in nature, wherein rehabilitation is proposed to play a critical role in improving or returning an individual’s functioning to its maximum. The selected health conditions do not present an exhaustive list, as additional health conditions will be added to the Package of Interventions for Rehabilitation in the future.^1^ For each of the health conditions, as a first step in developing the package, technical working groups comprised clinical and research experts who identified high-quality clinical practice guidelines and extracted relevant information, such as the actual recommendations and associated level of evidence. The reason for focusing on clinical practice guidelines at this stage of the process rather than other forms of evidence is because they are derived from rigorous systematic reviews of the best available research and practice experience and involve high-quality methodology during guideline development. Clinical practice guidelines also primarily focus on the effectiveness of interventions, provide a framework for clinical decision-making by producing recommendations to optimize patient care, and contain information regarding the benefits and harms of alternative care options predicated on evidence and value judgments. Technical working groups received methodological support by members of the WHO Rehabilitation Programme.

Of the selected health conditions, autism spectrum disorder (ASD) and intellectual disability (ID) are the focus of this report. Both conditions are neurodevelopmental disorders in the 5^th^ edition of the Diagnostic and Statistical Manual for Mental Disorders (DSM-5)^5^ and the 11^th^ edition of the International Classification of Disease (ICD-11).^6^ ASD is a disorder with onset in the early developmental period characterized by deficits in social communication and the presence of restricted, repetitive patterns of behavior.^5^ ID is defined by deficits in both intellectual and adaptive functioning during the developmental period, and is specified by unique codes based on the severity of adaptive functioning deficits, ranging from mild to profound impairment.^5,6^ By definition, individuals in the mild range may live fairly independent lives, while individuals in the profound range typically require constant supervision. We combined the findings for ASD and ID in this report because these disorders often co-occur, and similar aspects of adaptive functioning are impacted. Clinicians and researchers who work with individuals with one of these conditions also tend to work with individuals diagnosed with the other condition. Although the exact prevalence of ASD in people with ID is difficult to determine based on the literature, recent estimates indicate that ID is present in approximately 30-40% of individuals with ASD.^7,8^ Many people with ASD or ID have associated comorbid conditions, such as challenging behavior, mental health concerns, or associated health issues,^9,10^ which often tend to be the focus of intervention.

A vast array of medical and behavioral interventions have been developed and tested for different treatment targets and subpopulations of individuals with ASD, with varying degrees of reported rigor and efficacy.^11,12^ Heterogeneity in presentation, as well as unique needs for individuals across developmental trajectories and age spans, lead to few treatments for core symptoms and many treatments for co-occurring problems.^9^ For ID, the literature on management often includes general (non-individualized) principles.^13^ In addition, information on ID is often parsed based on specific genetic conditions with which it is associated.^14^ Due to the heterogeneity and varying severity levels of ID, approaches to treatment and rehabilitative services are varied.

The purpose of this report is to summarize the results of the search (including the extraction) and the analysis of rigorously vetted published clinical practice guidelines conducted by the technical working groups for rehabilitation in ASD and ID. The aims are to: (1) detail the search strategies employed to identify relevant clinical practice guidelines, (2) describe the screening process for determining eligible guidelines to be included in the Package of Interventions for Rehabilitation, (3) review the final selection of guidelines and corresponding recommendations, (4) present the recommendations categorized by type and target for each diagnostic group as well as by level of evidence, and (5) discuss research-to-practice gaps for future guideline development.

## Methods

High-quality clinical practice guidelines were sought based on a systematic search to identify relevant interventions for rehabilitation. These guidelines were selected based on specific criteria developed by the WHO Rehabilitation Programme.^1^

### Search Strategy

First, an extensive literature search was performed by each of the technical working groups for ASD and ID in six academic databases, one online search engine, 10 guideline databases, and seven professional rehabilitation society websites (see Table 1). The following general search criteria were used: “health condition (i.e., ASD or ID)” AND “rehabilitation” AND “guideline” (see Table S1 for full electronic search strategy of academic databases), restricted to 2008-2019 (search performed in January 2019), and limited to the English language.

**Table 1.**
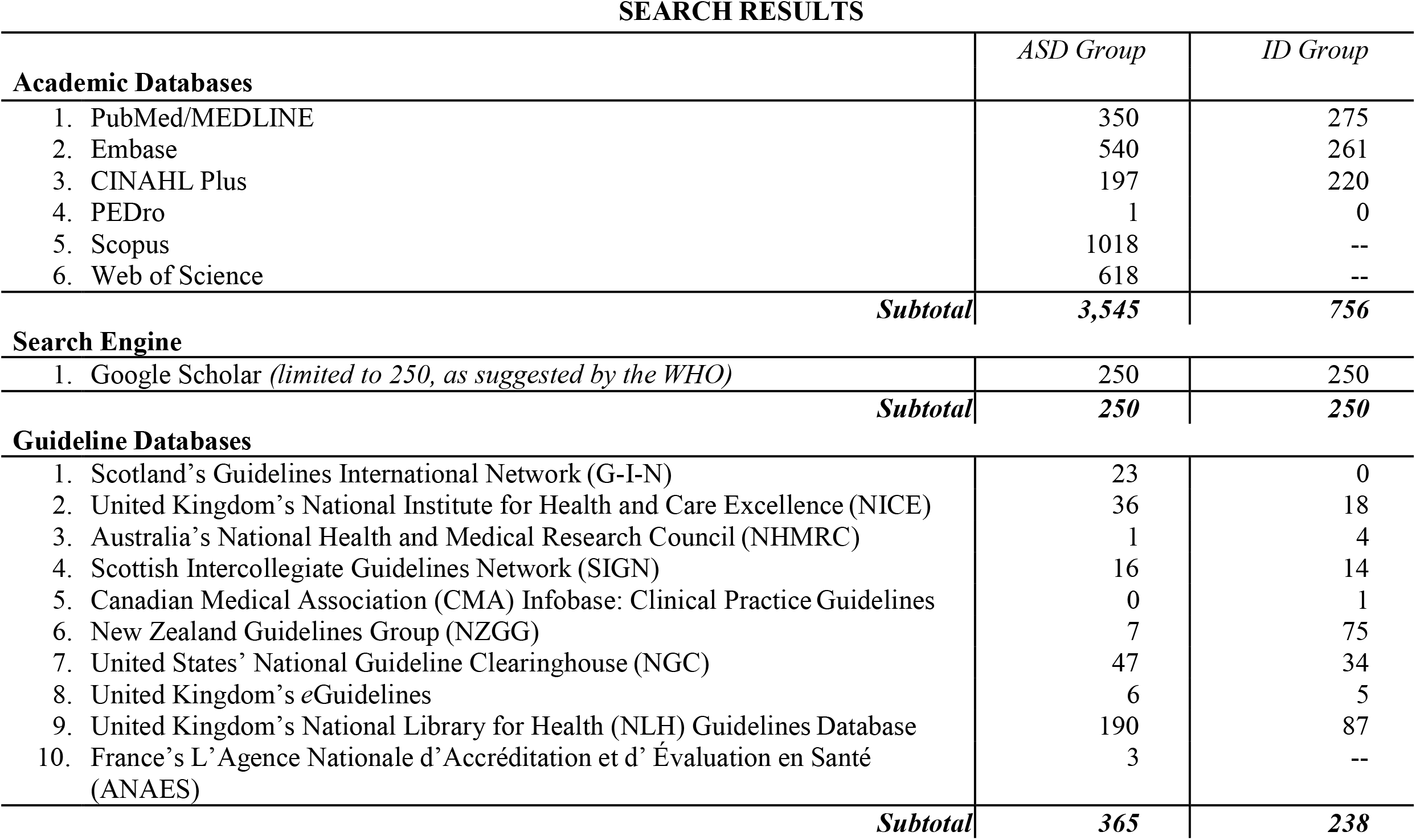
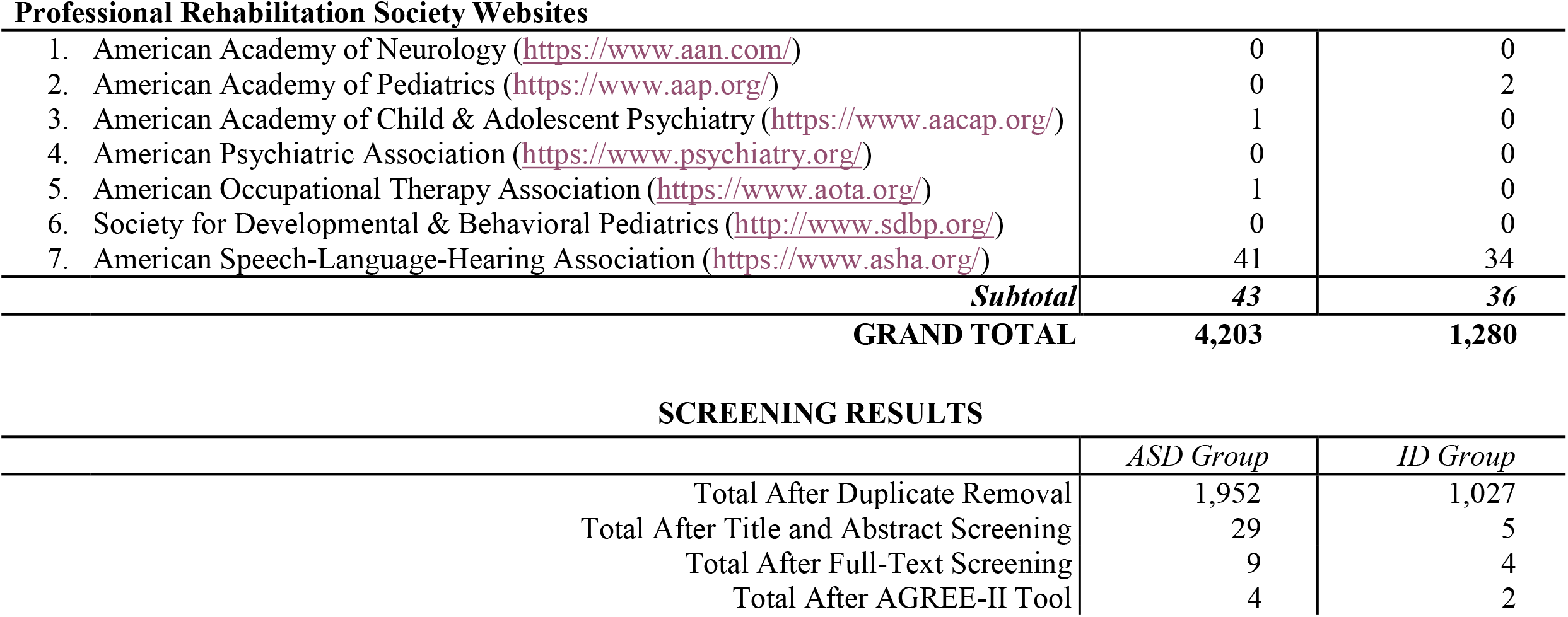
Results from the literature search and screening process; ‘--’ indicates the search was not performed.

| **SEARCH RESULTS** |  |  |
| --- | --- | --- |
|  | *ASD Group* | *ID Group* |
| **Academic Databases** |  |  |
| 1. PubMed/MEDLINE | 350 | 275 |
| 2. Embase | 540 | 261 |
| 3. CINAHL Plus | 197 | 220 |
| 4. PEDro | 1 | 0 |
| 5. Scopus | 1018 | -- |
| 6. Web of Science | 618 | -- |
| *Subtotal* | **3,545** | **756** |
| **Search Engine** |  |  |
| 1. Google Scholar (*limited to 250, as suggested by the WHO*) | 250 | 250 |
| *Subtotal* | **250** | **250** |
| **Guideline Databases** |  |  |
| 1. Scotland's Guidelines International Network (G-I-N) | 23 | 0 |
| 2. United Kingdom's National Institute for Health and Care Excellence (NICE) | 36 | 18 |
| 3. Australia's National Health and Medical Research Council (NHMRC) | 1 | 4 |
| 4. Scottish Intercollegiate Guidelines Network (SIGN) | 16 | 14 |
| 5. Canadian Medical Association (CMA) Infobase: Clinical Practice Guidelines | 0 | 1 |
| 6. New Zealand Guidelines Group (NZGG) | 7 | 75 |
| 7. United States' National Guideline Clearinghouse (NGC) | 47 | 34 |
| 8. United Kingdom's eGuidelines | 6 | 5 |
| 9. United Kingdom's National Library for Health (NLH) Guidelines Database | 190 | 87 |
| 10. France's L'Agence Nationale d'Accréditation et d'Évaluation en Santé (ANAES) | 3 | -- |
| *Subtotal* | **365** | **238** |
| **Professional Rehabilitation Society Websites** |  |  |
| 1. American Academy of Neurology (https://www.aan.com/) | 0 | 0 |
| 2. American Academy of Pediatrics (https://www.aap.org/) | 0 | 2 |
| 3. American Academy of Child & Adolescent Psychiatry (https://www.aacap.org/) | 1 | 0 |
| 4. American Psychiatric Association (https://www.psychiatry.org/) | 0 | 0 |
| 5. American Occupational Therapy Association (https://www.aota.org/) | 1 | 0 |
| 6. Society for Developmental & Behavioral Pediatrics (http://www.sdbp.org/) | 0 | 0 |
| 7. American Speech-Language-Hearing Association (https://www.asha.org/) | 41 | 34 |
| *Subtotal* | **43** | **36** |
| **GRAND TOTAL** | **4,203** | **1,280** |

**SCREENING RESULTS**
|  | *ASD Group* | *ID Group* |
| --- | --- | --- |
| Total After Duplicate Removal | 1,952 | 1,027 |
| Total After Title and Abstract Screening | 29 | 5 |
| Total After Full-Text Screening | 9 | 4 |
| Total After AGREE-II Tool | 4 | 2 |

### Identification of Suitable Guidelines

#### Title and Abstract Screening

Next, two members of each of the technical working groups independently screened the titles and abstracts of the articles retrieved from the search process. Articles were excluded for the following reasons: (1) not a guideline, (2) not about rehabilitation, (3) older than 10 years, or (4) developed for health conditions other than ASD or ID. The working group members then compared their decisions and discussed any discrepancies with a third member to reach consensus. In cases in which consensus could not be reached, the technical working group consulted with the WHO Rehabilitation Programme methods expert. Once the guidelines for inclusion were agreed upon, the full-text versions were screened.

#### Full-Text Screening

The same process for the title and abstract screening was applied during full-text screening, along with three additional exclusion criteria: (1) the presence of a conflict of interest (financial or non-financial), (2) the absence of information on the strength of the recommendation, or (3) did not pass the Appraisal of Guidelines for Research and Evaluation (AGREE-II)^15^ instrument criteria, which was applied to all guidelines that passed the first six exclusion criteria (items 1-4 in title and abstract screening and 1-2 in full-text screening). Three members of each working group independently determined AGREE-II ratings to assess the quality of each clinical practice guideline. All items were scored by each member, but only nine AGREE-II items (4, 7, 8, 10, 12, 13, 15, 22, 23) were specifically used for the selection of guidelines as pre-determined by the WHO Rehabilitation Programme.^1^ If the rating of any item differed by more than two points among the three members, the score was discussed to reach consensus. In cases in which agreement could not be reached, the groups consulted the WHO Rehabilitation Programme methods expert. Guidelines were excluded if the sum of the mean scores of the selected items was less than 45, or if the mean score of the three researchers on items 4, 8, 12 or 22 was below three.

### Data Extraction from Selected Guidelines

Technical working groups extracted specific information outlined by the WHO Rehabilitation Programme. Since the goal of this report is to describe the current recommendations from vetted guidelines that exist for ASD and ID, only the recommendations from the guidelines and their corresponding level of evidence are reported here. For the purposes of this manuscript, the reported level of evidence was categorized into one of two categories: ‘based on empirical evidence’ or ‘based on expert opinion’. The category empirical evidence consists of varying levels of evidence, including RCTs, meta-analyses, systematic reviews, case-control or cohort studies, and non-analytic studies (e.g., case reports, case series). We combined all levels of evidence into this empirical evidence category because each guideline used its own grading system for prescribing level of evidence. Conversely, expert opinion, the term used in the guidelines, conveys that members of the guideline development group reached clinical consensus to provide recommendations in cases in which there was insufficient empirical data to be prescribed a level of evidence. Additional classifiers were also included specifically for this report, to help summarize and describe the target focus of each recommendation by the following categories: ‘diagnostic group’ (ASD, ID), ‘general age group’ (children, young people, adults), ‘type of service’ (e.g., assessment, intervention), ‘target of service’ (what the recommendation was intended to help with), and ‘valence’ (do recommend, do not recommend, insufficient evidence to recommend). Recommendations could be classified into more than one category for both type of service and target of service. To identify the type of service and target of service, three working group members reviewed each recommendation and agreed upon categorizations through a three-step process: first the three members typed the specific word(s) used in the actual recommendation into the applicable table column, second the team members grouped the words/concepts from step one that were similar into overarching categories, and third the team members confirmed accuracy and consistency of decision-making for all categorizations applied to each recommendation in step two.

## Results

### Literature Search and Screening Results

A broad overview (flow chart in Figure 1) as well as a more detailed record (Table 1) characterize the process involved to obtain the resulting guidelines from the literature search for each group (ASD, ID).

**Figure 1.**
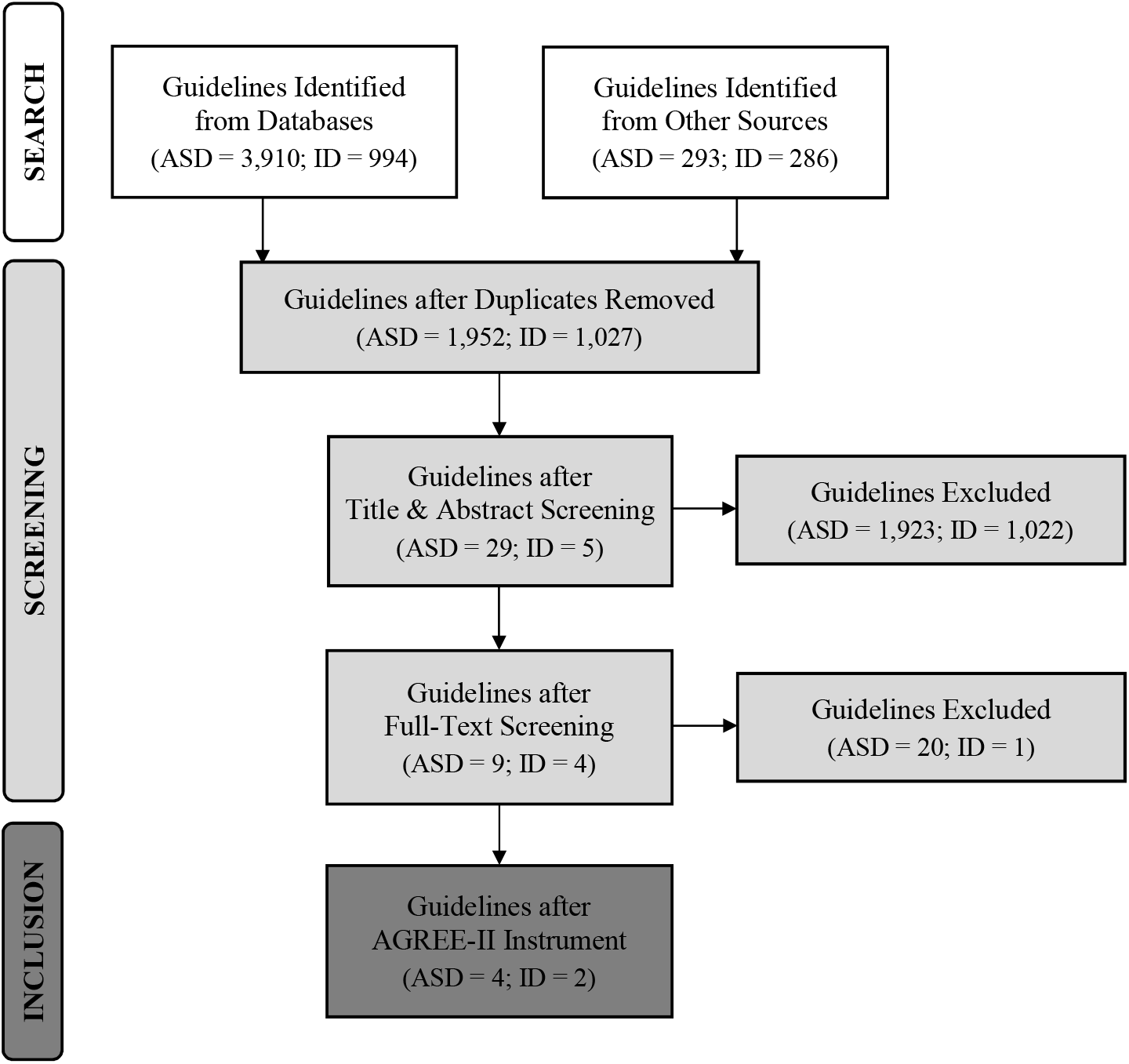
Flow chart of the results from the literature search and screening process.

#### ASD Guidelines

The literature search resulted in a grand total of 4,203 ASD articles across academic databases (n=3,545), search engines (n=250), guideline databases (n=365), and professional websites (n=43). After duplicate articles were removed, a total of 1,952 articles remained. Title and abstract screening reduced the number of articles to 29 guidelines, and full-text screening to nine guidelines. After applying the AGREE-II instrument (Table 2), four guidelines met the criteria^16-19^ and their descriptions are provided (Table 3). Two guidelines focused on treatment and support of youth,^17,18^ one targeted diagnosis and treatment in adults,^19^ and one concentrated on assessment, diagnosis, and treatment at all ages.^16^

**Table 2.** Results from the Appraisal of Guidelines for Research and Evaluation (AGREE-II) instrument for the six included guidelines are provided, with numbers representing the average of three reviewers’ scores. The nine items highlighted in gray were used for the selection of guidelines as pre-determined by the WHO Rehabilitation Programme, and guidelines were included if the sum of the mean scores of the selected items was at least 45.

| **AGREE-II Domain** | **AGREE-II Items** | **SIGN**<br>**145**<br>**(ASD)** | **KCE**<br>**233**<br>**(ASD)** | **NICE**<br>**CG170**<br>**(ASD)** | **NICE**<br>**CG142**<br>**(ASD)** | **NICE**<br>**NG11**<br>**(ID)** | **NICE**<br>**NG54**<br>**(ID)** |
| --- | --- | --- | --- | --- | --- | --- | --- |
| **1 Scope & Purpose** | **Item 1** | 6 | 7 | 7 | 7 | 7 | 7 |
|  | **Item 2** | 6 | 7 | 7 | 7 | 7 | 7 |
|  | **Item 3** | 5 | 7 | 7 | 7 | 7 | 7 |
| **2 Stakeholder Involvement** | **Item 4** | 6 | 6 | 7 | 6 | 6 | 6 |
|  | **Item 5** | 5 | 7 | 6 | 6 | 6 | 7 |
|  | **Item 6** | 7 | 6 | 7 | 7 | 6 | 7 |
| **3 Rigor of Development** | **Item 7** | 6 | 7 | 7 | 7 | 7 | 7 |
|  | **Item 8** | 3 | 6 | 6 | 6 | 6 | 7 |
|  | **Item 9** | 5 | 7 | 7 | 6 | 7 | 7 |
|  | **Item 10** | 2 | 7 | 5 | 5 | 5 | 6 |
|  | **Item 11** | 7 | 7 | 7 | 7 | 7 | 7 |
|  | **Item 12** | 5 | 6 | 7 | 7 | 7 | 7 |
|  | **Item 13** | 6 | 7 | 6 | 6 | 6 | 4 |
|  | **Item 14** | 6 | 4 | 5 | 7 | 7 | 7 |
| **4 Clarity & Presentation** | **Item 15** | 6 | 6 | 6 | 6 | 6 | 6 |
|  | **Item 16** | 6 | 6 | 6 | 6 | 6 | 6 |
|  | **Item 17** | 7 | 7 | 7 | 7 | 7 | 7 |
| **5 Applicability** | **Item 18** | 2 | 5 | 2 | 1 | 2 | 2 |
|  | **Item 19** | 6 | 4 | 6 | 3 | 5 | 4 |
|  | **Item 20** | 4 | 2 | 7 | 6 | 7 | 6 |
|  | **Item 21** | 4 | 1 | 4 | 3 | 1 | 1 |
| **6 Editorial Independence** | **Item 22** | 6 | 7 | 5 | 6 | 5 | 5 |
|  | **Item 23** | 6 | 6 | 5 | 7 | 6 | 6 |
|  | **Average of Highlighted Items** | **46** | **58** | **54** | **55** | **54** | **53** |

**Table 3.** Descriptions of each guideline consisting of the organization, guideline number, title, reference, year published, and scope.

| Organization (Guideline #) | Title [Reference] | Year | Scope (Text Taken From Each Guideline) |
| --- | --- | --- | --- |
| Scottish Intercollegiate Guidelines Network (SIGN 145) | Assessment, diagnosis and interventions for autism spectrum disorders: A national clinical guideline [17] | 2016 | This guideline provides recommendations based on current evidence for best practice in the assessment, diagnosis and interventions for children, young people, adults and older adults with ASD. It includes screening and surveillance, diagnosis and assessment, clinical interventions and service provision, as well as recommendations for research and audit. |
| Belgian Health Care Knowledge Centre (KCE 233) | Management of autism in children and young people: A good clinical practice guideline [18] | 2014 | This guideline provides recommendations based on current scientific evidence for treatment and support of children and adolescents with autism and their family. ASD is a complex condition. Therefore, addressing six domains, as defined by NICE, allows to discriminate outcomes. Per domain specific scores may be described. The objective of treatment is to improve outcomes specific to a particular domain of ASD, and thereby improving the overall outcome for the child or adolescent with ASD. Caregivers are encouraged to interpret these recommendations in the context of the individual situation and to take into account the values and preferences of the children and adolescents and of their families. |
| National Institute for Health and Care Excellence (NICE CG170) | Autism spectrum disorder in under 19s: Support and management [19] | 2013 | This guideline covers children and young people with autism spectrum disorder (across the full range of intellectual ability) from birth until their 19th birthday. It covers the different ways that health and social care professionals can provide support, treatment and help for children and young people with autism, and their families and carers, from the early years through to their transition into young adult life. |
| National Institute for Health and Care Excellence (NICE CG142) | Autism spectrum disorder in adults: Diagnosis and management [20] | 2012 | This guideline covers diagnosing and managing suspected or confirmed autism spectrum disorder (autism, Asperger's syndrome and atypical autism) in people aged 18 and over. It aims to improve access and engagement with interventions and services, and the experience of care, for people with autism. |
| National Institute for Health and Care Excellence (NICE NG11) | Challenging behaviour and learning disabilities: Prevention and interventions for people with learning disabilities whose behaviour challenges [21] | 2015 | This guideline covers interventions and support for children, young people and adults with a learning disability and behaviour that challenges. It highlights the importance of understanding the cause of behaviour that challenges, and performing thorough assessments so that steps can be taken to help people change their behaviour and improve their quality of life. The guideline also covers support and intervention for family members or carers. |
| National Institute for Health and Care Excellence (NICE NG54) | Mental health problems in people with learning disabilities: Prevention, assessment and management [22] | 2016 | This guideline covers preventing, assessing and managing mental health problems in people with learning disabilities in all settings (including health, social care, education, and forensic and criminal justice). It aims to improve assessment and support for mental health conditions, and help people with learning disabilities and their families and carers to be involved in their care. |

#### ID Guidelines

A grand total of 1,280 ID articles were found in the literature search across academic databases (n=756), search engines (n=250), guideline databases (n=238), and professional websites (n=36). After the removal of duplicate articles, 1,027 articles remained. Title and abstract screening reduced the number of articles to five guidelines, and full-text screening to four guidelines. Two guidelines^20,21^ met the AGREE-II instrument criteria (Table 2) for data extraction and are described (Table 3). Both guidelines focused on prevention and treatment in individuals of all ages, with one guideline targeting challenging behaviors^20^ and the other mental health problems.^21^

### Data Extraction Results

Data extraction results for level of evidence and the additional classifiers described in the Methods are provided in Table S2. For a visual breakdown of categories arranged by type of service and target of service, refer to Figures S1 and S2, respectively. Out of 524 (386 ASD; 138 ID) total recommendations, 52% (270) focused specifically on intervention (212 ASD; 58 ID), which is the focus of the Package of Interventions for Rehabilitation and therefore the focal point of this paper. The proportion of intervention recommendations categorized by each type of intervention and each target of intervention are presented in Figure 2A. For type of intervention, pharmacological was the primary category, accounting for 29% of both ASD and ID intervention recommendations, followed by biomedical and psychosocial for ASD (23% and 21%, respectively) and behavioral and psychological for ID (14% for each). For target of intervention, coexisting conditions were the primary category, encompassing 56% of ASD and 93% of ID intervention recommendations, while adaptive functioning was only 11% of ASD and 7% of ID intervention recommendations. Core symptoms of ASD were targeted in 26% of recommendations. Regarding the valence of intervention recommendations, only 67% of ASD compared to 100% of ID recommendations were categorized with a positive valence, meaning the intervention was recommended (as opposed to specifically not recommended or lacking sufficient evidence to provide a recommendation).

**Figure 2A.**
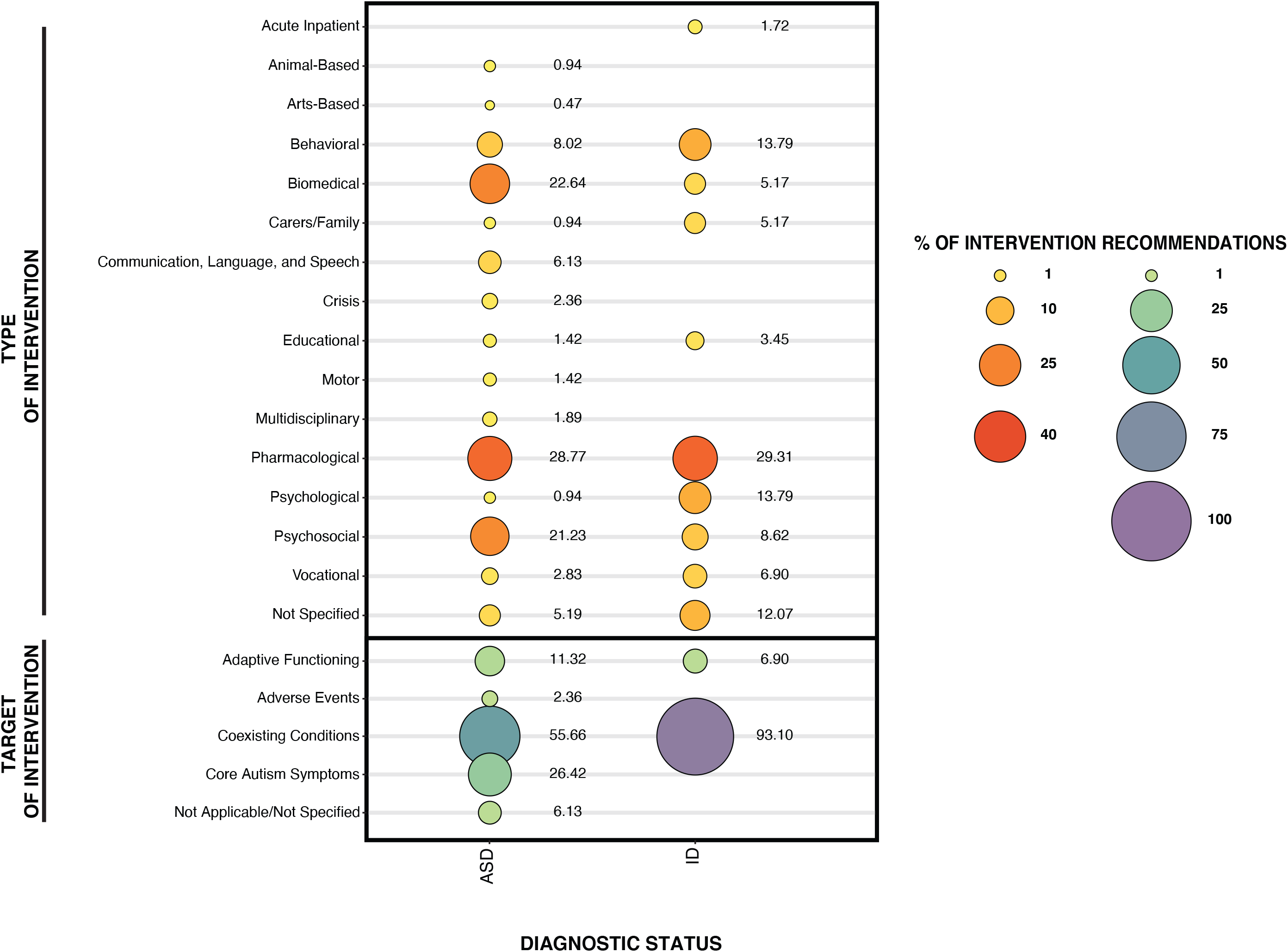
Percentage of intervention recommendations organized by ‘Type of Intervention’ and ‘Target of Intervention’ for autism spectrum disorder (ASD) and intellectual disability (ID).

For both disorders, many of the intervention recommendations were based on expert opinion, with only 17% (n=36) of ASD and 81% (n=47) of ID recommendations based on empirical evidence. The percentage of intervention recommendations based on empirical evidence are categorized by type of intervention and target of intervention (Figure 2B). Of these evidence-based intervention recommendations, the same trends remained, with pharmacological as the primary type of intervention accounting for 36% of both ASD and ID intervention recommendations, followed by biomedical and psychosocial for ASD (19% for each) and behavioral and psychological for ID (17% for each). For target of intervention, coexisting conditions remained as the primary category with evidence-based intervention recommendations, accounting for 39% of ASD and 100% of ID recommendations. Most of the ASD and ID recommendations had positive valence, 61% and 100%, respectively. Regarding age group, 12% were about children only (n=4 ASD, 6 ID), 22% were about children and young people, (n=18 ASD, 0 ID), 48% were about children, young people, and adults (n=0 ASD, 40 ID), and 18% were about adults only (n=14 ASD, 1 ID).

**Figure 2B.**
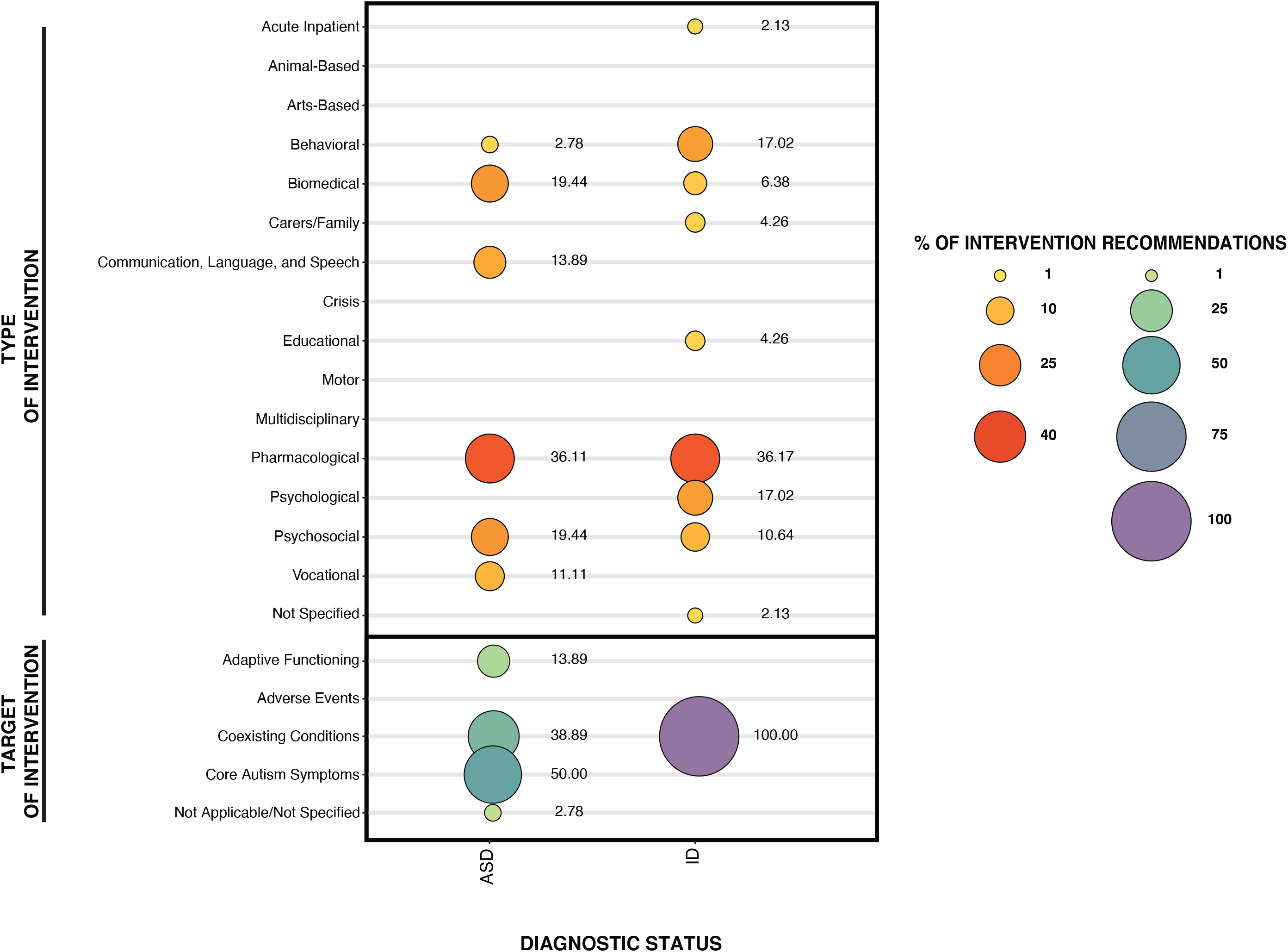
Percentage of intervention recommendations based on empirical evidence organized by ‘Type of Intervention’ and ‘Target of Intervention’ for autism spectrum disorder (ASD) and intellectual disability (ID).

## Discussion

This report included an extensive review of the literature to identify and extract information from clinical practice guidelines for ASD and ID for potential inclusion in the WHO Rehabilitation Programme’s Package of Interventions for Rehabilitation. The overall goal of rehabilitation is to achieve optimal functioning, which is a result of the interaction amongst a person’s health condition, environment, and personal context. For people with ASD and ID, achieving an optimal level of functioning may lead to increased independence and ability to participate in their environment (e.g., via improvements in self-determination, social and vocational fulfillment, community inclusion, meaningful participation). The WHO Rehabilitation Programme took this information into account when deciding to include ASD and ID in the Package of Interventions for Rehabilitation. Very few guidelines (six) regarding rehabilitation for ASD and ID met criteria for inclusion, all of which were produced by government agencies. Across the six guidelines, there were 524 recommendations in total, 270 of which related to intervention, and only 83 of which were based on empirical evidence. Although intervention recommendations are the focus of this report, it is noteworthy that 45% of ASD and 58% of ID recommendations were not categorized as intervention (i.e., assessment, general care, residential care, or care related to transition periods). Within the category of intervention, the majority of recommendations primarily comprised biomedical, pharmacological, and psychosocial interventions for ASD, and behavioral, pharmacological, and psychological interventions for ID, with pharmacological as the largest category for both.

With respect to the quality of the recommendations, the level of detail varied across recommendations and guidelines. Recommendations were generally classifiable in relation to the type of provider who may be trained to carry them out (e.g., medical provider, mental health provider, behavior therapist) but had limited information regarding other important details, such as skill level or training necessary for a practitioner to feasibly implement the recommended interventions. Additionally, the specificity of the recommendations in terms of the context or particular subpopulations for which they could be applicable was insufficient. Details regarding dosage, intensity, and duration of recommendations were usually not provided. In addition, few recommendations mentioned specific measures or programs; rather, the majority used general intervention categories instead (e.g., classroom-based interventions, interventions to support communication, sleep interventions). In addition, and perhaps most important to highlight, the majority of recommendations were based on expert opinion rather than empirical evidence (e.g., randomized control trials).

Regarding the intervention recommendations that were based on empirical evidence, some generalizations can be made for each diagnostic group. For individuals with ASD, it was recommended to address: (1) challenging behavior with antipsychotic medication only after other interventions have been unsuccessful, (2) mental health concerns with Selective Serotonin Reuptake Inhibitors or cognitive behavioral therapy (group or individual), (3) social communication and interaction problems with social learning programs (group or individual), (4) social isolation with structured leisure activity programs (group or individual), (5) employment needs with individual supported employment programs, (7) sleep difficulties with melatonin only after behavioral interventions have not worked, and (8) increased independence, communication, and social skills with applied behavioral analysis techniques.

For individuals with ID, recommendations based on empirical evidence were in reference to (1) challenging behavior, (2) mental health problems, and (3) sleep problems. For challenging behavior, it was generally stated that individuals with ID benefit from structured daily routines, cognitive behavioral therapy (specifically for anger management and depression), parent-training groups, exposure and relaxation therapy (for anxiety and phobias), and antipsychotic medication (only if other behavioral interventions have not been effective or if it is a high-risk problem behavior). Regarding the use of psychotropic medications, it was recommended these medications be prescribed by specialists, such as psychiatrists or neurodevelopmental pediatricians. It was also recommended that proactive strategies be used as much as possible to limit the use of reactive strategies. In the rare case that restrictive interventions must be used, they should be conducted ethically, documented, and reviewed regularly to determine whether continued use is warranted. For children ages 3-5 who are at-risk for or already have emerging challenging behavior, classroom-based interventions were recommended. Lastly, for sleep problems, behavioral interventions were recommended as the first approach, and then medications as the last resort (with melatonin a preferred choice).

Coexisting conditions were the main target of intervention recommendations for both ASD and ID. This is partly because the ID guidelines concentrated specifically on challenging behavior and mental health problems; however, it underscores the fact that there exist few intervention guidelines targeting the core symptoms of ASD or ID. For both ASD and ID, lack of sensitive outcome measures and rigorously controlled studies have been a stumbling block for research targeting core symptoms.^22-24^ One exception to this may be social skills, as there were a number of interventions targeting speech/language/communication and social skills deficits in the guidelines. However, there remain few or no intervention recommendations for the other core symptoms of ASD (e.g., restricted and repetitive patterns of behavior) or ID (low intellectual and adaptive functioning). This is a noteworthy gap because interventions for core symptoms could improve individuals’ quality of life and ability to participate in the community. In general, the lack of focus on adaptive behavior as an intervention target was surprising. Further examination of data contributing to this should focus on whether studies targeting such skills failed or were not implemented in the first place. Due to the major challenges of heterogeneity and treatment research that affect these disorders, an individually-tailored intervention approach is often used based on individual factors; recommendations based on empirical evidence are still being developed, as improved emerging approaches, treatment research designs, and outcome measures continue to enhance the field.^25-28^ This may be why so many of the recommendations are for specific approaches to a particular problem or point of care, rather than for a holistic approach to intervention.

Although some guidelines focused on specific sub-populations (e.g., children, adults) or on specific targets (e.g., challenging behaviors, mental health problems), a critical finding of this review revealed the need to consider more refined sub-populations within (or across) ASD and ID. Specifically, since the service needs of any given individual are fluid and dynamic, one gap to be filled is to provide recommendations that can be tailored according to individual needs. This involves contextualizing recommendations on age, severity of core symptoms, cognitive ability, language level, adaptive functioning, support needs, comorbid symptom profile, sociocultural context, or known etiologies/syndromes. Within both ASD and ID, there is significant heterogeneity in symptom presentation, across the populations and over time, and therefore diversity of rehabilitative needs, which negates the notion of a ‘one size fits all’ guideline. Furthermore, functional deficits of either condition may indicate that recommendations developed for one disorder may actually be appropriate for the other, and it is a goal of the Package of Interventions For Rehabilitation to have recommendations that are designed to cut across various conditions and be grouped by functioning domains.^1^ This is particularly relevant for ASD and ID given their likelihood of co-occurrence, the prevalence of similar comorbidities, and patterns of adaptive functioning deficits.

Although the term ‘rehabilitation’ suggests that a loss of functioning or a significant deviation from baseline occurred – which is the case in the majority of the other conditions selected for the Package of Interventions for Rehabilitation (e.g., spinal cord injury, stroke, traumatic brain injury) – the WHO defines rehabilitation as a set of interventions designed to optimize functioning and reduce disability in individuals with health conditions in interaction with their environment. Interventions for rehabilitation can be delivered from the time of onset of a disease or disorder, including those with onset in early childhood. Thus, the terms typically used in the developmental disability field to describe approaches designed to develop skills that may never have been learned in the first place (‘habilitation’, ‘treatment’, or ‘intervention’) can be interpreted as interventions optimizing functioning, and thus, as interventions for rehabilitation from a WHO perspective.

Importantly, the developmental progression of skills for people with ASD (in the area of socialization, at a minimum) and ID is expected to occur on a slower trajectory than for individuals who are typically-developing. Therefore, for interventions in ASD and ID, the goal is not to restore the individual to a previous level of functioning but rather to improve already-existing skills as much as possible in order to maximize functioning. Services traditionally conceptualized as rehabilitative, including speech therapy, occupational therapy, and physical therapy, have long been among the most frequently employed in targeting function in both ASD and ID.^29^ The WHO employs a broader definition of rehabilitation which allows for the consideration of other types of interventions (e.g., behavioral therapy and potentially psychopharmacologic interventions). Neither condition has Food and Drug Administration or otherwise-approved treatments for improving core symptoms, and to date, traditionally-considered rehabilitative services have also not proven to be efficacious in improving core symptoms.^30^ Therefore, the current ASD and ID guidelines presented here have mostly been focused on providing general recommendations for treating conditions and behavior that frequently co-occur with these disorders, but are not universal or specific to either diagnosis.

As part of the Package of Interventions for Rehabilitation initiative, the next step involves the convening development groups to work on the following tasks: (1) selection of interventions from those identified from the clinical practice guidelines to be included in the Package of Interventions for Rehabilitation, (2) identification of essential interventions that are missing in the selected guidelines, (3) description of required resources (workforce, assistive technologies, equipment, and consumables) and assignment to service delivery platforms for the interventions that are selected. For the selection of the interventions, the specific focus involves taking into account the available evidence, the number of people who will benefit from having access to interventions, and a good cost-benefit ratio. This process will include consideration of the inherent structural issues related to extrapolating guidelines from high-income countries to low- and middle-income countries when working toward a global health movement.

### Study Limitations

Although we consider this an extensive review of the guideline literature, there are still some limitations. It is possible that we may have had an incomplete retrieval of clinical practice guidelines that contain relevant intervention recommendations in ASD and ID due to the restrictions on language (English-only guidelines), publication year (guidelines published in the last ten years), and selection of search databases. In addition, we had to make inferences about the types and targets of service contained within each recommendation, as this information was not always explicitly stated in the guidelines. Lastly, our decision to combine recommendations supported by each level of empirical evidence into one group (in order to compare findings across guidelines) limits the ability to delineate recommendations based on higher-level evidence from those based on lower-level evidence.

## Conclusions

In sum, the diagnoses alone do not directly translate into a common set of treatment targets. The Package of Interventions for Rehabilitation will include the sets of interventions targeting aspects of functioning that are relevant to people with the different diagnoses. This begs the question of the utility of guidelines based on diagnosis alone versus recommendations that can be tailored according to the behavioral needs of each individual. We argue that guidelines related to the provision of interventions for rehabilitation should be organized and implemented based on behavior or functioning rather than diagnosis.

## Data Availability

This is a systematic review that included the following 6 guidelines (with links):
National Institute for Health and Care Excellence (NICE). Autism spectrum disorder in under 19s: support and management. Clinical guideline CG170. NICE. https://www.nice.org.uk/guidance/cg170. Published 2013. Accessed.
National Institute for Health and Care Excellence (NICE). Autism spectrum disorder in adults: diagnosis and management. Clinical guideline CG142. NICE. https://www.nice.org.uk/guidance/cg142. Published 2012. Accessed.
National Institute for Health and Care Excellence (NICE). Challenging behaviour and learning disabilities: prevention and interventions for people with learning disabilities whose behaviour challenges. NICE guideline NG11. NICE. https://www.nice.org.uk/guidance/ng11. Published 2015. Accessed.
National Institute for Health and Care Excellence (NICE). Mental health problems in people with learning disabilities: prevention, assessment and management. NICE guideline NG54. NICE. https://www.nice.org.uk/guidance/ng54. Published 2016. Accessed.
Scottish Intercollegiate Guidelines Network (SIGN). Assessment, diagnosis and interventions for autism spectrum disorders: A national clinical guideline. SIGN publication no. 145. SIGN. http://www.sign.ac.uk. Published 2016. Accessed.
Veereman G, Holdt Henningsen K, Eyssen M, et al. Management of autism in children and young people: a good clinical practice guideline. KCE reports 233. Belgian Health Care Knowledge Centre (KCE). https://kce.fgov.be/report/233. Published 2014. Accessed.

## Abbreviations

AGREE-II: Appraisal of Guidelines for Research and Evaluation
ASD: Autism Spectrum Disorder
ID: Intellectual Disability
WHO: World Health Organization

