## Supplementary material for "Systematic Review: Recommendations for Rehabilitation in ASD and ID from Clinical Practice Guidelines": Figures S1 and S2

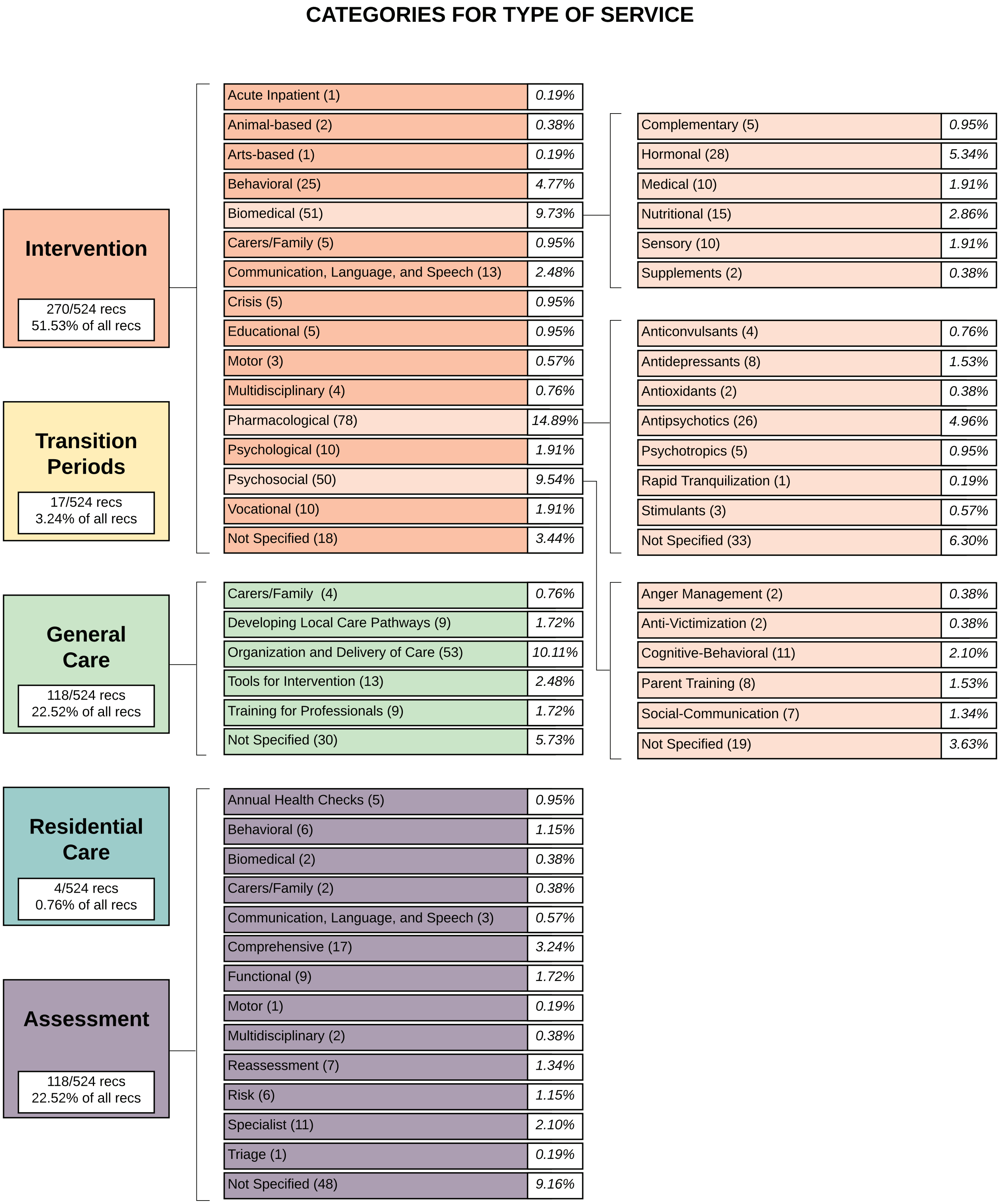


*Figure S1*. Percentage of all recommendations categorized by type of service for autism spectrum disorder (ASD) and intellectual disability (ID).

### CATEGORIES FOR TARGET OF SERVICE

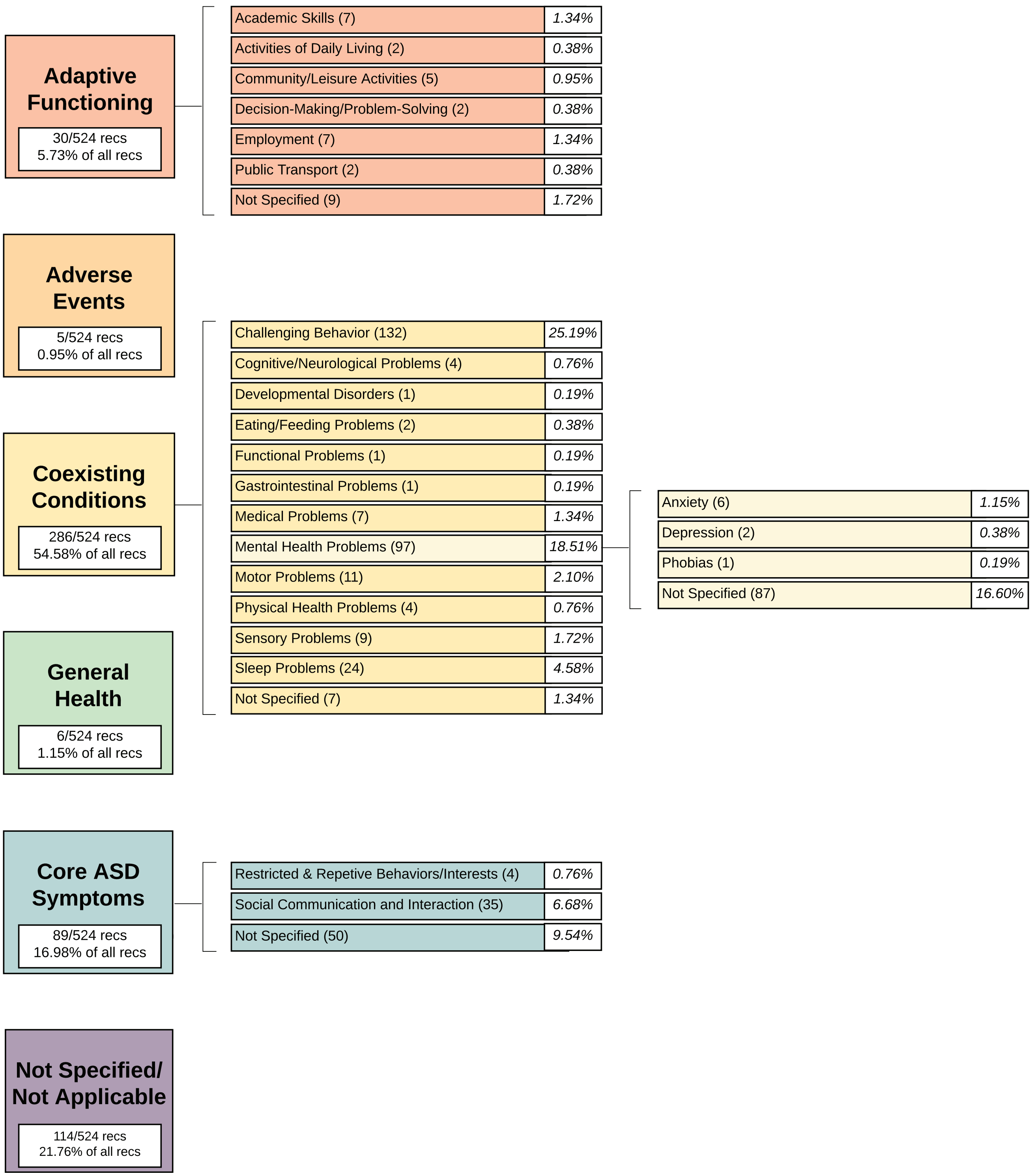


*Note: No subgroups or subcategories were used for these broad categories.

*Figure S2.* Percentage of all recommendations categorized by target of service for autism spectrum disorder (ASD) and intellectual disability (ID).
